# Short term Biological Variation of Differential Count in Healthy Subjects in a South Asian Population

**DOI:** 10.1101/2020.04.23.20076372

**Authors:** Arvind Kumar Gupta, K. Arathi, Sakshi Garg Tayal, Anoushika Mehan, Rishabh Sahay, Utpal Kumar, Michael Leonard Anthony, Neha Singh, Harish Chandra, Nilotpal Chowdhury

## Abstract

**Introduction:** Estimates of Within-Subject and between subject biological variation in South Asia are sorely lacking. Therefore, we attempted to estimate the short-term estimates of the same for Differential Counts (DC).

**Methods:** The study was conducted on twenty eight healthy volunteers (15 males and 13 females). After morning collection of blood in K3-EDTA vials analysis was conducted in triplicate on the Sysmex XN-1000 analyzer for six consecutive days. The Within subject, between subject and analytical coefficient of variation of the DC was calculated from the results by nested repeated measures ANOVA after outlier exclusion. The Reference change values (RCV) were also calculated.

**Results:** The within-subject variation for Eosinophil Count in our group from South Asia was greater than the published European and American studies. The between-subject variation in monocytes and basophils were also increased. Males and females showed similar biological variation for DC. The Within-subject variation of other parameters (Neutrophils, Lymphocytes, Monocytes and Basophils) were similar or showed only mild differences to the published studies.

**Conclusion:** The markedly different within-subject variation for Eosinophil counts suggest that the RCV for DC in South Asians need to be different from the published data in order to have clinical relevance. The Within-subject variation values of the other parameters seem transportable from the published European and American studies, but small differences found mean that further regional estimates need to be found for robust evidence of the same.

## INTRODUCTION

Within-subject biological variation of laboratory parameters has been recognized to be important for determining changes in the healthy state of an individual, as well as in setting analytical quality standards of a clinical pathology laboratory^1–3^. Knowledge of the expected biological variation can help determine if a person is deteriorating significantly in some parameters on serial follow up, especially with knowledge of the Reference Change Value (RCV). The RCV serves to provide the acceptable limits by which the value of an analyte can change in a person maintaining a “normal” homeostatic state and is calculated from the within-subject coefficient of variation (CV_I_). Also, acceptable imprecision and bias of automated instruments are best determined by knowledge of the CV_I_ and between-subject coefficient of variation (CV_G_). In recognition of the importance of CV_I_ and CV_G_, a database of the CV_I_ and CV_G_ of various analytes curated from published studies is being maintained by the European Federation of Laboratory Medicine (EFLM) at http://biologicalvariation.eu.

However, studies to estimate the CV_I_ and CV_G_ are difficult to conduct and needs relatively complicated statistical methods. Therefore, studies on the biological variation of many analytes have been performed in just a few areas in Europe and North America^4–10^. There have been especially few studies in an Asian population^11^. For the South Asian population, only one study has been performed for common hematological parameters, but none for the white blood cell differential counts (DC). Knowledge of the biological variation in such large unstudied populations is important since we do not know whether the European and North American estimates would remain equally valid in such a setting. Therefore, we decided to estimate the within and between subject coefficients of variation for the DC, comprising of the Neutrophil Count (NC), Lymphocyte Count (LC), Eosinophil Count (EC), Monocyte Count (MC) and Basophil Count (BC).

## MATERIALS AND METHODS

This study is part of a project approved by the Institutional Ethics Committee of a tertiary care institute in North India, part of which has already been published^12^. This was done on twenty-eight healthy volunteers, after a comprehensive physical and pathological check-up. There were 15 males (aging between 24 years to 41 years) and 13 females (aging between 21 years to 52 years).

The sample collection and analysis procedure for this study has been detailed previously^12^. In brief, samples were collected from all volunteers for six consecutive days between 9.00 AM and 10.00 AM. The analysis was conducted on a quality-controlled Sysmex XN-1000 blood analyzer each day. The Sysmex XN 1000 analyzer estimates the Differential counts by flow-cytometry using semiconductor laser after red cell lysis and permeabilization in a dedicated channel separate from the Nucleated red-cell channel, resulting in accurate counts^13,14^. The analysis started within four hours of collection and the values for NC, LC, EC, MC and BC extracted from the analyzer.

### Statistical analysis

Within subject and between subject heterogeneity of variance was detected by the Cochran test. Within-subject and between-subject outliers were detected by the Dixon test, and Normality assessed by the Shapiro-Wilk test. Then a nested repeated measures Analysis of Variance using linear mixed effects (LME) model was fitted and the examined graphically by residual plots and normal plot of the residuals. Potential outliers at the replicate level were detected on the residual. After examination and exclusion of outliers, the LME was run again and examined to ensure an adequate fit. The LME was then repeated on subsets comprising of only males and only females. Details of the replicates/subjects excluded from analysis with reasons are given in Table 1.

**Table 1:** Details of outliers removed during analysis. Outliers were screened by Dixon test for Samples within subject and mean of measurements between subjects. At the replicate level, outliers were screened from the residual plot by checking the z value of the standardized residual. Homogeneity of variance was detected by the Cochran test on residuals for both within-subject and between subject variance.

|  | Deleted after screening for outliers and further examination |  |  | Deleted after testing for homogeneity of variance |  | No of replicates in study |
| --- | --- | --- | --- | --- | --- | --- |
|  | replicate | sample | subject | sample | subject |  |
| Neutrophil count | 1 | 0 | 0 | 0 | 0 | 503 |
| Lymphocyte count | 1 | 0 | 0 | 0 | 1 | 485 |
| Eosinophil Count | 0 | 0 | 1 | 0 | 2 | 450 |
| Monocyte count | 0 | 0 | 1 | 0 | 1 | 468 |
| Basophil count | 0 | 0 | 0 | 0 | 0 | 504 |

The within-subject coefficient of variation (CV_I_), analytical coefficient of variation (CV_A_), and the between-subject coefficient of variation (CV_G_) was obtained by the SD-ANOVA method. The 95% Confidence Interval (C.I.) for the Various Coefficients of Variation were obtained by a parametric bootstrap with 5000 resamplings using the percentile method. A steady state for the experiment was verified by a quantile regression on the median. Finally, the Index of Individuality (II, calculated as 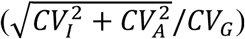, RCV (calculated as 1.96 *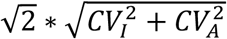 and the desirable specification for acceptable imprecision (<0.5 ∗ *CV*_*l*_) and acceptable bias 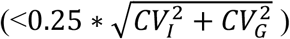 were calculated.

The sample size of 28 was sufficient to result in a power greater than 99% in detecting a CV_I_. This was also estimated to result in a precise 95% C.I. width less than 1/3rd of the absolute CV_I_ assuming that the CV_A_ was less than a half of the CV_I_.^15^

The R statistical environment, version 3.6.2, with the help of lme4, boot and outliers packages were used for the statistical analysis.^16–19^

## RESULTS

The results of the CV_I_,CV_A_ and CV_G_, along with RCV, acceptable imprecision, bias, and II are given in Table 2. The plots of the ranges of measurements are given in Figure 1. The comparison of the CV_I_ and CV_G_ of published studies taken http://biologicalvariation.eu is given in the Supplementary file. The CV_I_ and CV_G_ of LC, MC and BC are similar to published high quality studies, but substantial differences between the published studies and the present study were found for EC. For NC, the observed estimates agree with some biological variation studies^4,5^. The CV_G_ of the present studies differs with the database with respect to Monocytes and especially Basophils.

**Table 2:** Showing the Within-Subject Variation (CV_I_), Between Subject Variation (CV_G_), Analytical Variation (CV_A_), Reference Change Value (RCV), and estimates for acceptable imprecision (I%), bias (B%), and Index of Individuality (II) for all subjects, males and females.

|  | **CVI** | **CVG** | **CVA** | **RCV** | **I%** | **B%** | **II** |
| --- | --- | --- | --- | --- | --- | --- | --- |
| **Overall** |  |  |  |  |  |  |  |
| Neutrophil | 15.8 [13.3 ,18.6] | 29 [20.6 ,38.1] | 1.7 [1.5 ,1.9] | 44 | 7.9 | 8.3 | 0.5 |
| Lymphocyte | 10.6 [9.1 ,12.3] | 24.6 [17.5 ,31.9] | 2.5 [2.2 ,2.8] | 30.1 | 5.3 | 6.7 | 0.4 |
| Eosinophil | 19.5 [15.2 ,26] | 59 [39.4 ,84.9] | 7 [5.6 ,9.3] | 57.6 | 9.8 | 15.5 | 0.4 |
| Monocyte | 13.2 [11.3 ,15.3] | 20.4 [14.3 ,27] | 5.4 [4.8 ,6.1] | 39.5 | 6.6 | 6.1 | 0.7 |
| Basophil | 10.7 [7.7 ,13.8] | 40.6 [28.6 ,54.5] | 18.2 [15.5 ,22] | 58.5 | 5.3 | 10.5 | 0.5 |
| **Male** |  |  |  |  |  |  |  |
| Neutrophil | 16 [12.6 ,20.4] | 33.1 [19.9 ,47.9] | 1.5 [1.3 ,1.9] | 44.4 | 8 | 9.2 | 0.5 |
| Lymphocyte | 12.1 [9.7 ,14.7] | 23.3 [14 ,32.5] | 2.5 [2.1 ,2.9] | 34.1 | 6 | 6.5 | 0.5 |
| Eosinophil | 18.9 [13.5 ,27.3] | 56.3 [32.5 ,90.3] | 7.8 [5.9 ,11.3] | 56.6 | 9.4 | 14.8 | 0.4 |
| Monocyte | 13.3 [10.6 ,16.1] | 18.7 [10.4 ,26.8] | 5.2 [4.5 ,6.1] | 39.5 | 6.6 | 5.7 | 0.8 |
| Basophil | 10.3 [5.9 ,14.8] | 42.3 [25.3 ,62.9] | 18.8 [15 ,24.3] | 59.6 | 5.2 | 10.9 | 0.5 |
| **Female** |  |  |  |  |  |  |  |
| Neutrophil | 15.6 [12.4 ,19.4] | 25.1 [13.9 ,36.9] | 1.9 [1.6 ,2.2] | 43.4 | 7.8 | 7.4 | 0.6 |
| Lymphocyte | 8.7 [6.7 ,10.9] | 26.5 [15.2 ,39] | 2.5 [2.1 ,3.1] | 25.1 | 4.3 | 7 | 0.3 |
| Eosinophil | 19.8 [13.5 ,31.7] | 60.3 [31.4 ,106.4] | 6.3 [4.5 ,9.7] | 57.5 | 9.9 | 15.9 | 0.3 |
| Monocyte | 13.1 [10.3 ,16.2] | 22.6 [12.5 ,32.6] | 5.6 [4.8 ,6.7] | 39.5 | 6.6 | 6.5 | 0.6 |
| Basophil | 11 [6.5 ,15.6] | 39.4 [22.9 ,60] | 17.5 [13.9 ,23.2] | 57.3 | 5.5 | 10.2 | 0.5 |

**Figure 1:**
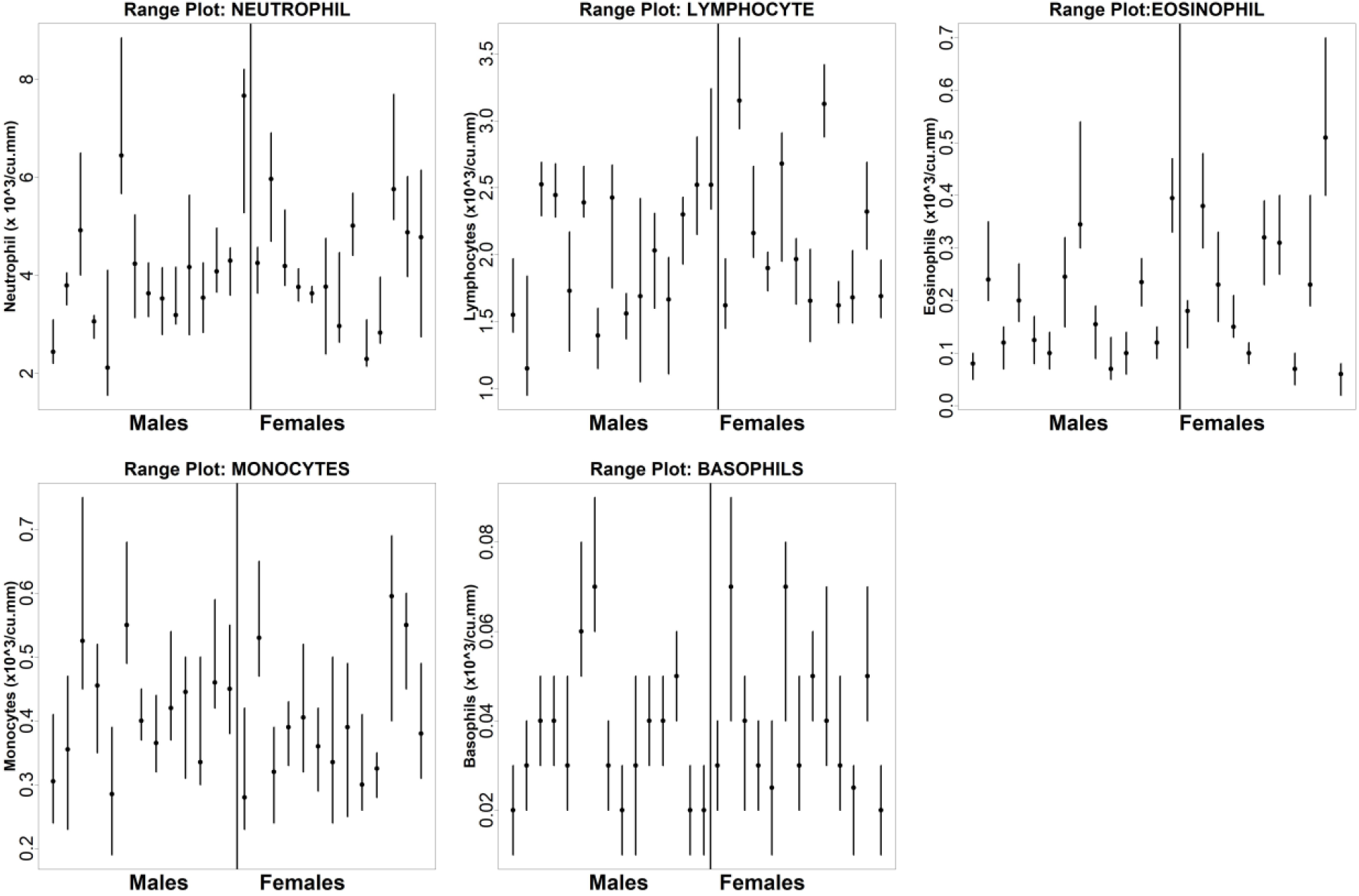
Range plot of the various parameters of the Differential Count.

## DISCUSSION

This is possibly the first study to report on the biological variation of the differential count parameters from South Asia. While the results of LC, MC and BC are compatible with the published Western studies, the differences in EC hint that separate biological variation values may be needed for making clinical decisions in Subcontinental India for differential counts. Even the milder differences found for NC suggest the need for further studies on biological variation in South Asia which are, sadly, lacking.

The RCV (possibly) more suitable for the South Asian population have been presented. The CV_I_ was far lower than the CV_G_ for all the Differential Count parameters (and reflected in the II values). This suggests that decisions based on the RCV may be more sensitive in detecting pathological events rather than waiting for a change in parameter values exceeding the reference interval limits. However, such usefulness can only be documented in clinical studies, and our Department is planning to initiate clinical studies to test whether addition of the CV_I_ related metrics could actually lead to a more sensitive and early diagnosis.

The CV_I_ of the Eosinophil count is first hematological parameter we encountered in our project^12^ differing in within-subject variability from Western studies. While racial/genetic differences may explain some of these differences, we feel that is more likely that continued exposure to allergens in the environment may be the primary reason for this increased variability. North India, and other areas of the Indian Subcontinent, have markedly increased air pollutants with values commonly reaching hazardous PM_2.5_ values^20,21^. This increased chronic dust exposure may serve as lung irritants, resulting in a background Type I immune response^22^. Also, this may serve as mild inflammatory mediators in the lung, aiding the mildly increased neutrophilic within-subject variability. Parasitic infestation may serve as an unlikely cause; but all participants were clinically healthy.

Biological variation data is also used for determining the quality standards of automated clinical pathology analyzers. For most of the analytes, the observed analytical precision met the desired standards. For Basophils, however, this was not matched, reflecting the difficulty in having a high precision in parameters with low cell counts. Also, since the majority of the parameters closely agreed with the published European studies, the analytical quality standards used in European laboratories should remain applicable for their Indian counterparts at least for the DC parameters.

While this was a part of a project which was previously published, the present results (of the DC) were delayed since there was difficulty in fitting the ANOVA models. This was due to marked heterogeneity of the variance for Eosinophil count in some volunteers compared to the others which had a limiting effect on the statistical analysis (which pre-supposes homogeneity of variances). We could only overcome this problem by completely deleting the EC results of three volunteers from the analysis by using strict outlier detection and exclusion protocol. To avoid completely deleting some volunteers from analysis, we had previously attempted to fit our statistical models differently, with weighted regression allowing for differing variances among subjects^23^. While the above approach worked well statistically, it was possibly less generalizable. Therefore, we decided to keep the present approach of strict outlier exclusion.

The present study is valid predominantly for short term estimates of biological variation. Longer term estimates of the biological variation have been suggested to differ from short term studies^5^; long term studies on hematological parameters are however limited by the difficulty in estimating reagent lot to lot variation and the effects of analytical changes due to possible routine calibration and validation of the equipment, and the impossibility of storing hematological samples in a stable state for a long period of time like serum samples. However, we do intend to carry out longer term studies in the future.

In conclusion, the present study publishes, possibly for the first time in South Asia) the short term within-subject and between subject variation for the differential count parameters. In doing so, we have demonstrated that there are differences in the CV_I_ in EC which possibly points out the need for having differing clinical RCVs for DC in South Asia, as also the need for more data on the Within-Subject and Between-Subject Variation from the same area.

## Data Availability

All data incorporated with this manuscript

